# Ecological Momentary Assessments of Trauma-Related Intrusive Memories: Potential Clinical Utility

**DOI:** 10.1101/2024.05.15.24307377

**Authors:** Yara Pollmann, Kevin J. Clancy, Quentin Devignes, Boyu Ren, Milissa Kaufman, Isabelle M. Rosso

## Abstract

As the global prevalence of trauma rises, there is a growing need for accessible and scalable treatments for trauma-related disorders like posttraumatic stress disorder (PTSD). Trauma-related intrusive memories (TR-IMs) are a central PTSD symptom and a target of exposure-based therapies, gold-standard treatments that are effective but resource-intensive. This study examined whether a brief ecological momentary assessment (EMA) protocol assessing the phenomenology of TR-IMs could reduce intrusion symptoms in trauma-exposed adults.

Participants (N=131) experiencing at least 2 TR-IMs per week related to a DSM-5 criterion A trauma completed a 2-week EMA protocol during which they reported on TR-IM properties three times per day, and on posttraumatic stress symptoms at the end of each day. Longitudinal symptom measurements were entered into linear mixed-effects models to test the effect of Time on TR-IMs.

Over the 2-week EMA protocol, intrusion symptom severity (cluster B scores) significantly declined (t = −2.78, p = 0.006), while other symptom cluster scores did not significantly change. Follow-up analyses demonstrated that this effect was specific to TR-IMs (t = −4.02, p < 0.001), and was not moderated by survey completion rate, total PTSD symptom severity, or ongoing treatment.

Our findings indicate that implementing an EMA protocol assessing intrusive memories could be an effective trauma intervention. Despite study limitations like its quasi-experimental design and absence of a control group, the specificity of findings to intrusive memories argues against a mere regression to the mean. Overall, an EMA approach could provide a cost-effective and scalable treatment option targeting intrusive memory symptoms.

## INTRODUCTION

The global occurrence of trauma is rising steeply, with a growing prevalence of war, displacement, natural disasters, terrorism, violence, and other humanitarian crises. It is estimated that over 70% of the world’s population has been exposed to a traumatic event in their lifetime [1]. The impact of trauma at the individual, community, and societal level is staggering – with data showcasing its association with increased risk of mental health disorders, low educational outcomes, and unemployment, exacting an annual economic toll of 3 billion in lost productivity in the United States alone [2–4]. Existing guidelines for gold-standard care for trauma-related psychiatric disorders, such as posttraumatic stress disorder (PTSD), list Cognitive Processing Therapy, Eye Movement Desensitization and Reprocessing, and Prolonged Exposure Therapy as the most empirically supported and recommended treatments [5]. However, barriers persist in trauma care, including high dropout rates and significant loss to follow-up undermining efficacy, shortage of trained therapists and limited in-person care options impeding accessibility, and sociocultural factors impacting acceptability [6–8]. Accordingly, there is a pressing need for innovative treatment approaches [9].

In addressing the need for novel therapies, there has been growing emphasis on targeting core, transdiagnostic symptoms that maintain distress and impairment, rather than solely focusing on categorical diagnoses [10]. This approach is particularly important considering the marked heterogeneity of PTSD and its associated symptom structure [11]. This complexity is compounded when considering trauma exposure more comprehensively, as it often leads to adverse mental health outcomes beyond PTSD [12]. For instance, trauma-exposed individuals may exhibit only one or a few posttraumatic stress symptoms but experience significant distress and/or impairment nonetheless [13]. Moreover, they may experience a constellation of symptoms meeting diagnostic criteria for other psychiatric disorders [14].

Intrusive re-experiencing of the traumatic event is prevalent among trauma-exposed individuals, most commonly manifesting as trauma-related intrusive memories (TR-IMs). TR-IMs are involuntary, emerging spontaneously and intruding on conscious thought processes. They manifest in a multitude of ways, notably in the form of images, but also as smells, sounds, tastes, bodily sensations, and emotional responses [10,15]. Crucially, TR-IMs are key predictors of the development, severity, and maintenance of PTSD [16] and other adverse post-traumatic mental health outcomes [12], making them a critical therapeutic target for trauma survivors.

Among the gold-standard treatment options for PTSD, prolonged exposure (PE) therapy has the strongest focus on TR-IMs. PE accomplishes this through fear extinction, whereby patients repeatedly focus on details of a traumatic experience in a safe environment until fear is reduced [17]. Despite its proven efficacy [18], PE and similar exposure therapies face significant obstacles, including initial symptom exacerbation, high dropout rates, and dissemination barriers due to its dependence on highly-trained clinicians and environmental support [6,7,19]. Written exposure therapy (WET), a newer PTSD treatment grounded in exposure therapy principles, was developed to address these barriers by offering a brief, accessible protocol that involves repeated writing of trauma narratives [20]. Research shows that WET is non-inferior to PE [21], underscoring the clinical utility of brief, less resource-intensive protocols. However, despite these advances, barriers such as reliance on clinician involvement and limited transdiagnostic utility persist, leaving a significant portion of trauma-exposed individuals with TR-IMs without adequate intervention.

In light of these considerations, we sought to examine the potential clinical utility of brief, periodic reporting of the occurrence and phenomenological properties of TR-IMs, which entails brief exposure to details of the traumatic event. Ecological momentary assessment (EMA), a real-time data collection tool that periodically samples participants’ experiences [22], has been used to track PTSD symptoms naturalistically. While prior research has examined the impact of EMA and other related momentary assessments on symptom change [23–29], no study has yet investigated the clinical utility of EMA in alleviating intrusive memory symptoms specifically. Through secondary analyses of an independent project utilizing EMAs of the phenomenological properties of TR-IMs in trauma-exposed adults, we aimed to determine whether EMA could offer a more accessible therapeutic option, potentially addressing the current limitations of exposure-based therapies. We hypothesized that completing an EMA protocol, during which participants reported on the phenomenological properties of their TR-IMs, would be associated with a reduction in their intrusive re-experiencing symptom severity over time.

## PATIENTS AND METHODS

### Participants

Trauma-exposed adults (N=202) were recruited via multiple sources as part of a larger project examining the neurobiological correlates of TR-IMs [30,31]. The study was approved by the Mass General Brigham Human Research Committee, and all participants provided written informed consent. Inclusion criteria consisted of a) 18-65 years of age, b) exposure to at least one DSM-5 Criterion A trauma, c) endorsement of at least two TR-IMs per week in the past month, d) English proficiency, and e) access to a “MetricWire” (MetricWire Inc, Kitc) application-compatible smartphone. Exclusion criteria were a) current psychotic disorder or manic mood episode, b) report of experiencing intrusions only as thoughts, as opposed to memories, c) completion of less than 70% of the daily surveys during the EMA period, and d) past month moderate-to-severe alcohol or substance use disorder. Of the 202 participants, 20 did not complete all study procedures, 5 met diagnostic criteria for psychosis or a substance use disorder, 37 completed an insufficient number of EMA surveys, and 1 participant did not endorse any TR-IMs during the survey period and was thus excluded from analyses. Importantly, the 37 participants who did not complete a sufficient number of EMA surveys did not differ from the final analyzed sample in any PTSD symptom severity (p’s > 0.274). Due to technical difficulties, 8 participants had survey periods extended beyond 2 weeks. Given our interest in the effect of Time for the analyses presented here, these participants were excluded, resulting in a final analyzed sample of n = 131. Participant demographics and clinical characteristics are presented in Table 1.

**Table 1.** Demographic and clinical characteristics. Means ± standard deviations or N (%).

| Total Sample (N=131) |  |
| --- | --- |
| Age (years) | 34 ± 11.1 |
| Sex |  |
| Female | 98 (75%) |
| Male | 33 (25%) |
| Gender |  |
| Woman | 84 (64%) |
| Man | 32 (25%) |
| Non-binary | 15 (11%) |
| Race/Ethnicity |  |
| Asian | 6 (5%) |
| Black | 8 (6%) |
| White | 88 (67%) |
| Hispanic | 6 (5%) |
| Other | 2 (2%) |
| Multiracial | 21 (16%) |
| Asian and White | 9 (43%) |
| Black and White | 5 (24%) |
| Middle Eastern North African and White | 3 (14%) |
| Other | 4 (19%) |
| LEC-5 Total | 13.1 ± 7.76 |
| PCL-5 Total | 48.15 ± 13.46 |
| PTSD Diagnosis | 100 (76%) |
| Number of completed TR-IM EMA surveys | 36 ± 3.77 |
| Number of completed PCL-5 EMA surveys | 11.49 ± 2.08 |

EMA: Ecological Momentary Assessment; LEC-5: Life Events Checklist for DSM-5; PCL-5: PTSD Checklist for DSM-5; PTSD: Posttraumatic Stress Disorder; TR-IM: Trauma-Related Intrusive Memory.

### Procedure

The study design consisted of two visits separated by a two-week EMA survey period. During Visit 1, participants provided written informed consent before initiating study procedures. They then completed self-report measures and received detailed instructions along with a guided demonstration of the EMA smartphone application. Over the subsequent two-week EMA period, participants received 5 daily surveys. The first survey probed sleep quality and dreams/nightmares. The following three surveys assessed the occurrence and phenomenological properties of experienced TR-IMs. These surveys included adapted items from the Autobiographical Memory Questionnaire (AMQ; [32]) to measure the vividness, visual features, sense of nowness, emotional intensity, fragmentation, and intrusiveness of TR-IMs, rated on a 0-4 Likert scale. The final survey assessed PTSD symptom severity over that past day (24 hours) using the Posttraumatic Stress Disorder Checklist (PCL-5). The survey schedule was personalized based on each participant’s sleep/wake times, and surveys were sent at semi-random times within 3-hour windows, with the initial 3-hour window beginning one hour before the participant’s typical wake time. Participants who completed at least 70% of all EMA surveys returned for Visit 2 to complete a clinical interview and neuroimaging.

### Measures

#### Life Events Checklist (LEC-5)

The LEC-5 [33] is a 17-item assessment of potentially traumatic events, reflecting a Criterion A trauma. The LEC-5 was used during Visit 1 to identify and orient participants to their Criterion A index trauma for EMA survey completion. In addition, total LEC-5 score was derived as an index of total lifetime trauma exposure for sample characterization (Table 1).

#### PTSD Checklist for DSM-5 (PCL-5)

The PCL-5 [34] was administered daily during the EMA period to assess PTSD symptoms. It is a 20-item self-report assessment of the 20 DSM-5 symptoms of PTSD. It is frequently used to monitor symptom change during and after treatment. Although typically administered to assess PTSD symptom severity over the past month, the PCL-5 items were adapted for our EMA protocol to query symptoms experienced over the past day (24 hours), consistent with prior work using daily adaptations of the PCL-5 [35,36]. Scores are summed into DSM-5 PTSD symptom clusters, consisting of intrusive re-experiencing (Cluster B), avoidance (Cluster C), negative alterations in mood and cognition (Cluster D), and hyperarousal (Cluster E) symptoms. Of particular relevance to this study, the intrusion symptom cluster consists of 5 symptoms: 1) trauma-related intrusive memories (TR-IMs), 2) trauma-related nightmares or distressing dreams, 3) flashbacks, and 4) emotional and 5) physical reactions to trauma reminders.

#### Clinician-Administered PTSD Scale for DSM-5 (CAPS-5)

During Visit 2, the CAPS-5 [37] was administered by doctoral-level clinicians to determine PTSD diagnosis and clinician-rated symptom severity, including a score for total PTSD symptom severity (CAPS-5 Total; Table 1).

### Statistical Analyses

To test the effect of Time on intrusion symptoms, repeated measures of daily PCL-5 Intrusive Re-experiencing (Cluster B) symptom scores over time were entered in a linear mixed effect model (LMM) using the lme4 package [38]. The model incorporated subject-specific random intercepts and slopes. The independent variable of Time reflects the timestamp of each completed PCL-5 survey in days relative to the beginning of the survey period. Significant models were re-run separately, with the total number of completed memory surveys, CAPS-5 PTSD symptom severity, and a binary variable (1, 0) for ongoing psychotherapy entered as moderators of the effect of Time. Age and sex did not demonstrate associations with intrusion symptoms (p’s > 0.589) and were therefore not included in the models; however, results were virtually identical when including age and sex as covariates (detailed below).

To ascertain specificity of the effects to Intrusion symptoms targeted in the EMA protocol, identical models were performed for other PTSD symptom clusters (C, D, E) and each individual intrusion symptom. Correction for multiple comparisons was performed using Bonferroni correction (0.05/4 symptom clusters = 0.0125; 0.05/5 intrusion symptoms = 0.010).

## RESULTS

The linear mixed effects model predicting PCL-5 Cluster B severity revealed a significant effect of Time on daily intrusion symptoms (beta = −0.06, t = −2.78, p = 0.006), such that participants demonstrated a decrease in intrusion symptom severity over the EMA period (Figure 1A-C). This effect was independent of the total number of completed memory surveys (p = 0.479), total CAPS-5 PTSD severity (p = 0.742), or ongoing treatment (p = 0.868). Additionally, this effect was virtually identical when controlling for Age and Sex (t = −2.79, p = 0.006).

**Figure 1.**
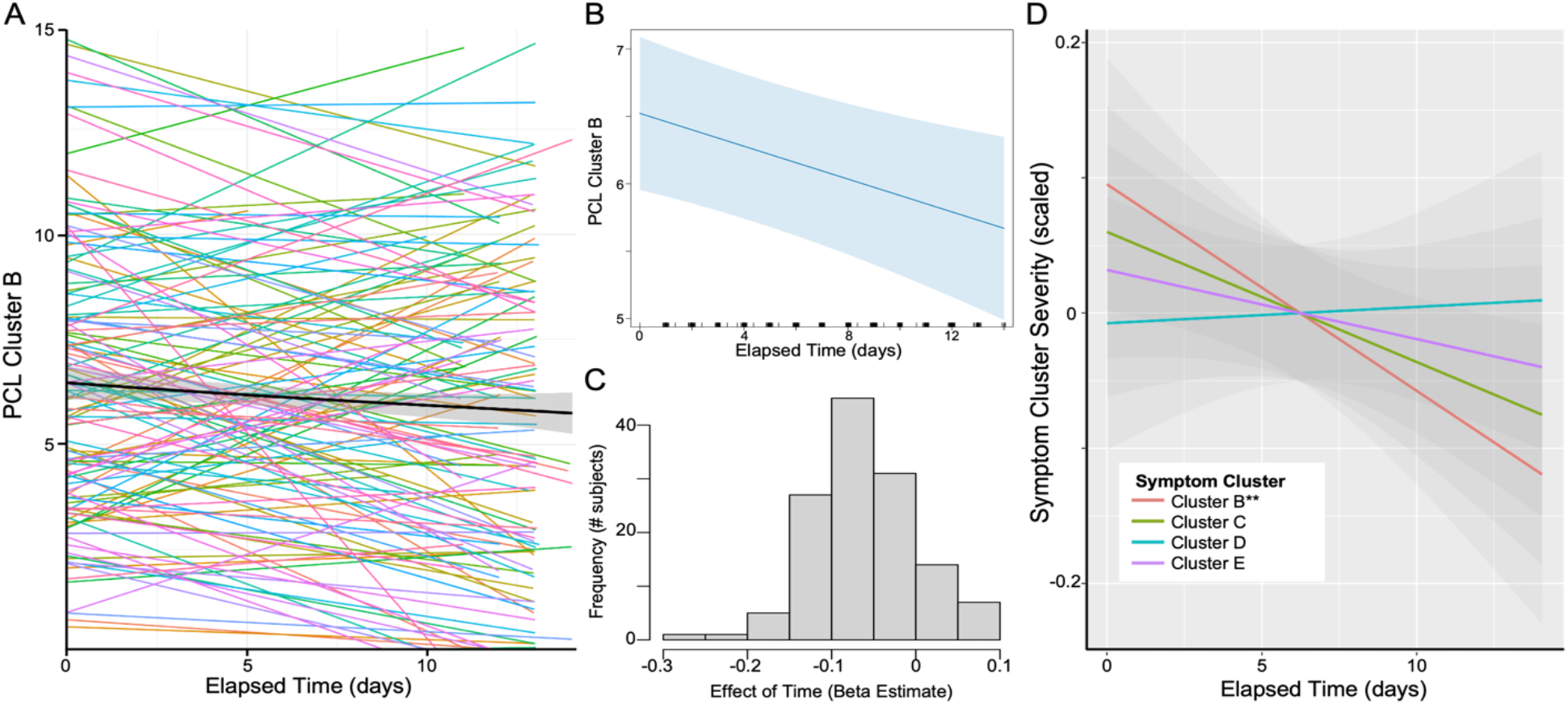
Effect of Time on Cluster B – Intrusive Re-experiencing Symptoms. **(A)** Individual lines represent subject-specific intercepts and slopes for the effect of time on PCL-5 Cluster B symptoms, with the group-level effect indicated by the black line, and further depicted in **(B). (C)** Distribution of the effect of time across participants, demonstrating that most participants showed a decrease in Cluster B symptoms over time. **(D)** Effect of time for each symptom cluster separately, demonstrating a significant effect on Cluster B score and no other symptom Cluster. To allow for uniform comparison across symptom clusters, cluster scores were scaled (mean-centered and divided by standard deviation). ** p<0.01.

Identical linear mixed models performed with other PCL-5 symptom clusters revealed no effect of Time for Cluster C (avoidance; t = −1.39, p = 0.167), Cluster D (mood/cognition; t = 0.16, p = 0.870), or Cluster E symptoms (hyperarousal; t = −0.77, p = 0.443; Figure 1D).

Specificity to individual TR-IM symptoms was examined through additional linear mixed models. A significant effect of Time was seen on trauma-related intrusive memories (beta = −0.02, t = −4.02, p < 0.001; Figure 2A-C), with no effect on nightmares (t = −1.42, p = 0.159), flashbacks (t = −1.48, p = 0.140), or physical reactivity to trauma reminders (t = −1.11, p = −0.269; Figure 2D). A weaker effect of Time was seen for emotional reactivity to trauma reminders, which did not survive correction for multiple comparisons (t = −2.47, p = 0.015). The effect of Time on intrusive memories was independent of the total number of memory surveys completed (p = 0.669), total CAPS-5 PTSD severity (p = 0.921), or ongoing treatment (p = 0.746). The effect on intrusive memories was virtually identical when controlling for Age and Sex (t = −4.02, p = 0.0001).

**Figure 2.**
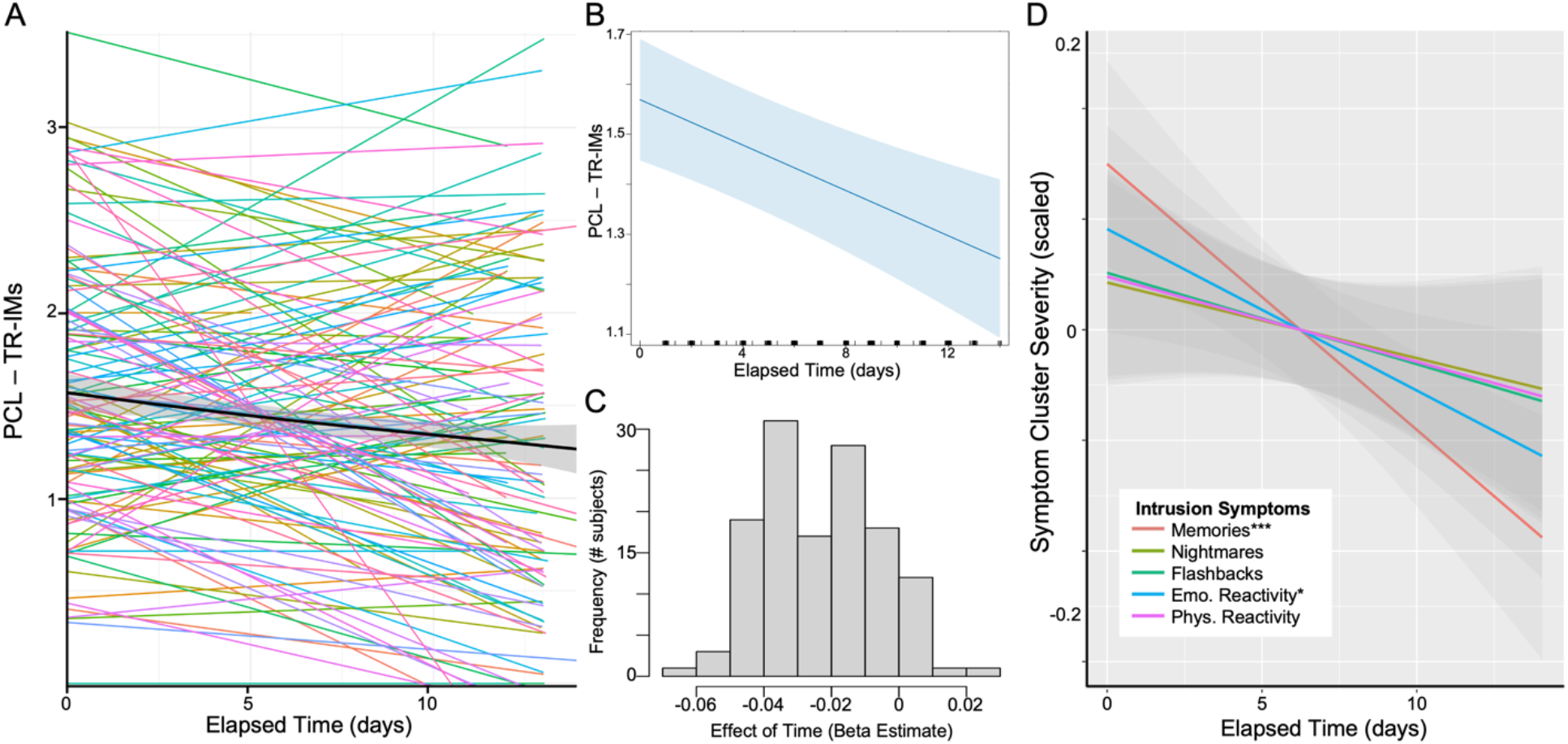
Effect of Time on Trauma-Related Intrusive Memories (TR-IMs). **(A)** Individual lines represent subject-specific intercepts and slopes for the effect of time on PCL-5 TR-IMs, with the group-level effect indicated by the black line, and further depicted in **(B). (C)** Distribution of the effect of time across participants, demonstrating that most participants showed a decrease in TR-IMs over time. **(D)** Effect of time for each Cluster B symptom separately, demonstrating a significant effect on TR-IMs and no other Cluster B symptom. To allow for comparison across symptoms, scores were scaled. * p<0.05, *** p<0.005.

## DISCUSSION

This study examined the potential clinical utility of EMA for intrusion symptoms in trauma-exposed adults. Our findings support the hypothesis that completing an EMA protocol assessing TR-IMs leads to a decrease in intrusion symptom severity. We observed a specific reduction in intrusion symptoms (Cluster B), while symptoms of avoidance (Cluster C), negative alterations in cognition and mood (Cluster D), and alterations in arousal and reactivity (Cluster E) did not significantly change over time. In addition, we found a significant reduction in the emotional reaction to memories. Specificity analyses revealed that the decrease in intrusion symptoms was specific to intrusive memories, consistent with the targeted nature of the EMA protocol. These findings provide preliminary evidence supporting the tenet that an EMA protocol assessing the phenomenology of trauma-related intrusive memories may reduce intrusion symptoms.

Our findings align with and build upon previous research demonstrating the therapeutic efficacy of self-monitoring for PTSD symptoms. Typically, EMA protocols are used as a data collection or monitoring tool, with less focus on their potential clinical effectiveness. Nonetheless, research has indicated that daily EMA surveys simply monitoring PTSD symptoms can lead to significant reductions in total PTSD symptom severity [24], setting the precedent that monitoring symptoms may result in their improvement. Consistent with this notion, other work has demonstrated specific improvements in intrusion symptoms following daily self-monitoring of traumatic intrusion frequency in PTSD patients [28]. This mirrors the specificity of our findings to intrusive memory symptoms when using EMA to assess trauma intrusions. Importantly, in our study, the EMA protocol goes beyond mere symptom monitoring by prompting participants to recall in detail the sensory, emotional, and cognitive properties of their TR-IMs [32], fostering greater engagement with and processing of these memories.

The detailed examination and reporting of TR-IMs is unique to our present EMA protocol and may more closely emulate elements of exposure-based therapies, potentially explaining the observed effect on intrusive memories. Previous research by Kleim and colleagues found that individuals with PTSD experienced diary-prompted voluntary trauma memories with a comparable sense of nowness and vividness as involuntary intrusive memories [25]. If this holds true, participants engaging with EMA-prompted memories may do so with a high level of engagement. This would potentially activate the ‘fear structure’ and facilitate emotional processing of trauma memories, which are key components of exposure therapies [17]. Moreover, the repeated imaginal reliving of memories throughout the two-week EMA period, facilitated by the daily completion of several surveys, may promote habituation - another key component of exposure therapies [39,40].

In keeping with the exposure framework, there are notable similarities between written exposure therapy (WET) and the present EMA protocol, namely in terms of exposure reporting technique, fewer exposure doses required, and shorter exposure duration. Our findings coalesce with parallel lines of research on the clinical efficacy of exposure-based approaches that are less intensive, shorter in duration, and do not rely on heavy clinician involvement, such as WET [21]. Both WET and the current protocol consist of daily reporting on trauma memories, and both offer promising avenues to reduce clinician demand and increase access to evidence-based psychotherapies. However, WET involves 5, 1-hour sessions consisting of a 30-minute writing period followed by 30 minutes of clinician guidance and direction. The current EMA protocol consists of three 2-minute surveys about the details of the traumatic memory throughout the day, suggesting greater accessibility and acceptability with this approach. As such, the present EMA protocol potentially offers an additional avenue to overcome barriers to exposure-based treatments, with a deeper focus on practicality, ease of use, and limited reliance on resources and personnel.

Regarding personnel demands, the current protocol showcased symptom improvement without clinician involvement. This parallels a growing body of literature demonstrating the efficacy of certain gold-standard treatments without clinician involvement, such as internet-based cognitive behavioral therapy [41,42]. However, there is limited evidence for such adaptations for exposure-based therapies, which rely on clinician involvement to varying degrees. Indeed, clinician involvement serves an important role in facilitating the appropriate engagement in clinical exposures. Nonetheless, recent work has demonstrated non-inferiority of an internet-based adaptation of WET that incorporates peer specialists instead of clinicians [43]. Addressing the barrier of clinician involvement is particularly important, given reports of clinician hesitation to engage in exposure-based therapies [44]. It has the additional potential benefit of promoting self-agency within patients [45–47], which may be particularly relevant for individuals with trauma-related disorders, given the loss of agency that often stems from trauma exposure [46]. Ultimately, a protocol that emulates elements of exposure-based therapies and can be implemented without the guidance of a clinician may be of particular value when considering these barriers and the therapeutic efficacy of self-agency.

Acknowledging this study’s limitations, the quasi-experimental design without a control group prompts caution in drawing definitive or causal conclusions. This is due to the data originating from an individual difference (within-subjects) study conducted for non-treatment purposes. Further research including a control group in a randomized clinical trial (RCT) is necessary to determine whether the observed intrusion symptom reductions were influenced by regression to the mean over time. Nonetheless, the specificity of these reductions to the intrusive memory symptoms that were targeted by the EMA protocol, along with a more stable trajectory of other symptoms, provide preliminary yet strong evidence against this explanation. Additionally, our results were robust to controlling for multiple factors, including ongoing psychotherapy, total clinician-rated PTSD severity, and number of surveys completed, which bolsters confidence in our findings.

Following validation of the preliminary clinical efficacy of an EMA protocol for TR-IMs through an RCT, there are several promising future directions. First, there are options to develop the EMA protocol into an ecological momentary intervention (EMI) framework by adapting or including additional prompts based on reported TR-IM properties, which may enhance clinical efficacy by increasing engagement and processing of TR-IMs. Additionally, there is potential for clinical utility across psychiatric disorders, given the role of intrusive memories in multiple different conditions [48–50]. Relatedly, there is a substantial need to address and treat subthreshold presentations of PTSD [13], as many evidence-based treatments are predicated on samples meeting full diagnostic criteria for PTSD [5]. By targeting TR-IMs, we can potentially reduce the risk of additional symptoms developing and progressing into full PTSD or other psychiatric disorders, given the centrality of intrusive symptoms [51,52]. Additionally, because this protocol was conducted entirely electronically via smartphone, it offers potential for widespread therapeutic use. This is bolstered by the adaptability, accessibility, cost-effectiveness, and cultural sensitivity that such technology-based mental health care offers [53]. While technology-based barriers exist, there is a rapid increase in technology use and access taking place in both developed and developing countries [54–56]. Incorporating community-based participatory approaches would help ensure the expansion of these technology-based approaches can maximally meet the different needs of those from different cultural and socioeconomic backgrounds.

In conclusion, these data provide preliminary evidence for the clinical utility of an EMA protocol assessing properties of TR-IMs, coalescing with recent advances in the expansion of exposure-based therapies. Through future RCTs, extensions of this EMA protocol have the potential to address the central and pervasive symptom of intrusive memories in an accessible, affordable, and culturally sensitive manner. Given that traumatic exposure is ballooning globally with the increase of natural disasters, war, violence, and terrorism, such scalable protocols may reduce the risk and burden of trauma-related psychiatric disorders globally. With the advance of technology and the growing increase of both technological literacy and access, protocols such as the one presented in the current study contribute to an influential movement towards technology-based care, advancing our ability to provide effective care for all.

## Data Availability

Data available upon reasonable request to the corresponding author (IMR), and following execution of a data use agreement.

## ACKNOWLEDGMENTS

We thank all participants who completed this study.

## AUTHOR CONTRIBUTIONS

**YP:** Conceptualization, Investigation, Data Curation, Writing-Original Draft.

**KJC:** Conceptualization, Formal Analysis, Visualization, Writing-Original Draft, Supervision.

**QD:** Data Curation, Supervision, Writing-Review and editing

**BR:** Data Curation, Formal Analysis, Writing-Review and editing

**MK:** Investigation, Supervision, Writing-Review and editing

**IMR:** Conceptualization, Investigation, Methodology, Project Administration, Funding Acquisition, Supervision, Writing-Review and editing,

## Notes

### Competing Interest Statement

The authors have declared no competing interest.

### Funding Statement

This research was funded by NIMH R01MH120400 (IMR).

### Author Declarations

Massachusetts General Brigham Human Research Committee

## REFERENCES

1. Benjet C, Bromet E, Karam EG, Kessler RC, McLaughlin KA, Ruscio AM, et al. The epidemiology of traumatic event exposure worldwide: results from the World Mental Health Survey Consortium. Psychol Med. 2016;46:327–343.

2. Kessler RC. Posttraumatic stress disorder: the burden to the individual and to society. J Clin Psychiatry. 2000;61 Suppl 5:4–12; discussion 13--14.

3. Noble NC, Merker JB, Webber TK, Ressler KJ, Seligowski AV. PTSD and depression severity are associated with cardiovascular disease symptoms in trauma-exposed women. Eur J Psychotraumatol. 2023;14:2234810.

4. Sowder KL, Knight LA, Fishalow J. Trauma exposure and health: A review of outcomes and pathways. J Aggress Maltreat Trauma. 2018;27:1041–1059.

5. VA/DoD TLG. VA/DoD Clinical Practice Guideline for Management of Posttraumatic Stress Disorder and Acute Stress Disorder. 2023. 2023.

6. Foa E, Gillihan S, Bryant R. Challenges and successes in dissemination of evidence-based treatments for posttraumatic stress lessons learned from prolonged exposure therapy for PTSD. Psychol Sci Public Interest. 2013;14:65–111.

7. Imel ZE, Laska K, Jakcupcak M, Simpson TL. Meta-analysis of dropout in treatments for post-traumatic stress disorder. J Consult Clin Psychol. 2013;81:394–404.

8. Kazlauskas E. Challenges for providing health care in traumatized populations: Barriers for PTSD treatments and the need for new developments. Glob Health Action. 2017;10:1322399.

9. Hoge CW, Chard KM. A window into the evolution of trauma-focused psychotherapies for posttraumatic stress disorder. JAMA. 2018;319:343–345.

10. Iyadurai L, Visser RM, Lau-Zhu A, Porcheret K, Horsch A, Holmes EA, et al. Intrusive memories of trauma: A target for research bridging cognitive science and its clinical application. Clin Psychol Rev. 2019;69:67–82.

11. Bryant RA, Galatzer-Levy I, Hadzi-Pavlovic D. The heterogeneity of posttraumatic stress disorder in DSM-5. JAMA Psychiatry. 2023;80:189–191.

12. Rossi FS, Nillni Y, Fox AB, Galovski TE. The association between lifetime trauma exposure typologies and mental health outcomes among veterans. Psychiatry Res. 2023;326:115321.

13. McLaughlin KA, Koenen KC, Friedman MJ, Ruscio AM, Karam EG, Shahly V, et al. Subthreshold posttraumatic stress disorder in the WHO world mental health surveys. Biol Psychiatry. 2015;77:375–384.

14. Kessler RC, Sonnega A, Bromet E, Hughes M, Nelson CB. Posttraumatic stress disorder in the National Comorbidity Survey. Arch Gen Psychiatry. 1995;52:1048–1060.

15. Ehlers A, Hackmann A, Michael T. Intrusive re-experiencing in post-traumatic stress disorder: phenomenology, theory, and therapy. Memory. 2004;12:403–415.

16. Ehlers A, Steil R. Maintenance of intrusive memories in posttraumatic stress disorder: A cognitive approach. Behav Cogn Psychother. 1995;23:217–249.

17. Rothbaum BO, Schwartz AC. Exposure therapy for posttraumatic stress disorder. Am J Psychother. 2002;56:59–75.

18. Powers MB, Halpern JM, Ferenschak MP, Gillihan SJ, Foa EB. A meta-analytic review of prolonged exposure for posttraumatic stress disorder. Clin Psychol Rev. 2010;30:635–641.

19. Larsen SE, Wiltsey Stirman S, Smith BN, Resick PA. Symptom exacerbations in trauma-focused treatments: Associations with treatment outcome and non-completion. Behav Res Ther. 2016;77:68–77.

20. Thompson-Hollands J, Marx BP, Sloan DM. Brief novel therapies for PTSD: Written Exposure Therapy. Current Treatment Options in Psychiatry. 2019;6:99–106.

21. Sloan DM, Marx BP, Acierno R, Messina M, Muzzy W, Gallagher MW, et al. Written exposure therapy vs prolonged exposure therapy in the treatment of posttraumatic stress disorder: A randomized clinical trial. JAMA Psychiatry. 2023;80:1093–1100.

22. Shiffman S, Stone AA, Hufford MR. Ecological momentary assessment. Annu Rev Clin Psychol. 2008;4:1–32.

23. Brown AJ, Bollini AM, Craighead LW, Astin MC, Norrholm SD, Bradley B. Self-monitoring of reexperiencing symptoms: A randomized trial. J Trauma Stress. 2014;27:519– 525.

24. Dewey D, McDonald MK, Brown WJ, Boyd SJ, Bunnell BE, Schuldberg D. The impact of ecological momentary assessment on posttraumatic stress symptom trajectory. Psychiatry Res. 2015;230:300–303.

25. Kleim B, Graham B, Bryant RA, Ehlers A. Capturing intrusive re-experiencing in trauma survivors’ daily lives using ecological momentary assessment. J Abnorm Psychol. 2013;122:998–1009.

26. Pedersen ER, Kaysen DL, Lindgren KP, Blayney J, Simpson TL. Impact of daily assessments on distress and PTSD symptoms in trauma-exposed women. J Interpers Violence. 2014;29:824–845.

27. Possemato K, Kaier E, Wade M, Lantinga LJ, Maisto SA, Ouimette P. Assessing daily fluctuations in posttraumatic stress disorder symptoms and substance use with Interactive Voice Response technology: protocol compliance and reactions. Psychol Serv. 2012;9:185– 196.

28. Reynolds M, Tarrier N. Monitoring of intrusions in post-traumatic stress disorder: A report of single case studies. Br J Med Psychol. 1996;69:371–379.

29. Tarrier N, Sommerfield C, Reynolds M, Pilgrim H. Symptom self-monitoring in the treatment of posttraumatic stress disorder. Behav Ther. 1999;30:597–605.

30. Clancy KJ, Devignes Q, Ren B, Pollmann Y, Nielsen SR, Howell K, et al. Spatiotemporal dynamics of hippocampal-cortical networks underlying the unique phenomenological properties of trauma-related intrusive memories. Mol Psychiatry. 2024. March 7, 2024. 10.1038/s41380-024-02486-9.

31. Devignes Q, Ren B, Clancy KJ, Howell K, Pollmann Y, Martinez-Sanchez L, et al. Trauma-related intrusive memories and anterior hippocampus structural covariance: an ecological momentary assessment study in posttraumatic stress disorder. Transl Psychiatry. 2024;14:74.

32. Rubin DC, Schrauf RW, Greenberg DL. Belief and recollection of autobiographical memories. Mem Cognit. 2003;31:887–901.

33. Gray MJ, Litz BT, Hsu JL, Lombardo TW. Psychometric properties of the life events checklist. Assessment. 2004;11:330–341.

34. Blevins CA, Weathers FW, Davis MT, Witte TK, Domino JL. The Posttraumatic Stress Disorder Checklist for DSM-5 (PCL-5): Development and initial psychometric evaluation. J Trauma Stress. 2015;28:489–498.

35. Alting van Geusau VVP, Mulder JD, Matthijssen SJMA. Predicting outcome in an intensive outpatient PTSD Treatment program using daily measures. J Clin Med Res. 2021;10:4152.

36. Vogel L, Koller G, Ehring T. The relationship between posttraumatic stress symptoms and craving in patients with substance use disorder attending detoxification. Drug Alcohol Depend. 2021;223:108709.

37. Weathers FW, Bovin MJ, Lee DJ, Sloan DM, Schnurr PP, Kaloupek DG, et al. The Clinician-Administered PTSD Scale for DSM-5 (CAPS-5): Development and initial psychometric evaluation in military veterans. Psychol Assess. 2018;30:383–395.

38. Bates D, Mächler M, Bolker B, Walker S. Fitting linear mixed-effects models using lme4. ArXiv E-Prints. 2014;arXi:1406.

39. Foa E, Kozak M. Foa EB, Kozak MJ. Emotional processing of fear. Exposure to corrective information. Psychol Bull 99: 20-35. Psychological Bulletin. 1986;99:20–35.

40. Jaycox L, Foa E, Morral A. Influence of emotional engagement and habituation on exposure therapy for PTSD. Journal of Consulting and Clinical Psychology, 66, 185-192. Journal of Consulting and Clinical Psychology. 1998;66:185–192.

41. Karyotaki E, Efthimiou O, Miguel C, Bermpohl FMG, Furukawa TA, Cuijpers P, et al. Internet-based cognitive behavioral therapy for depression: A systematic review and individual patient data network meta-analysis. JAMA Psychiatry. 2021;78:361–371.

42. Rosso IM, Killgore WDS, Olson EA, Webb CA, Fukunaga R, Auerbach RP, et al. Internet-based cognitive behavior therapy for major depressive disorder: A randomized controlled trial. Depress Anxiety. 2017;34:236–245.

43. McLean CP, Malek N, Straud CL. A pilot randomized controlled trial of online written exposure therapy delivered by peer coaches to veterans with posttraumatic stress disorder. J Trauma Stress. 2024. February 13, 2024. 10.1002/jts.23020.

44. Moses K, Gonsalvez CJ, Meade T. Barriers to the use of exposure therapy by psychologists treating anxiety, obsessive-compulsive disorder, and posttraumatic stress disorder in an Australian sample. J Clin Psychol. 2023;79:1156–1165.

45. Acke E, De Smet MM, Van Nieuwenhove K, Meganck R. The nature of client agency prior to therapy: A qualitative study on clients’ narratives. Psychol Belg. 2022;62:17–28.

46. Ataria Y. Sense of ownership and sense of agency during trauma. Phenomenol Cognitive Sci. 2015;14:199–212.

47. Yao X, Dong B, Ji W. Formulation and Clients’ Agency in Cognitive Behavioral Therapy. Frontiers in Psychology. 2022;13:810437.

48. Bisby JA, Brewin CR, Leitz JR, Valerie Curran H. Acute effects of alcohol on the development of intrusive memories. Psychopharmacology. 2009;204:655–666.

49. Brewin CR, Gregory JD, Lipton M, Burgess N. Intrusive images in psychological disorders: characteristics, neural mechanisms, and treatment implications. Psychol Rev. 2010;117:210–232.

50. Payne A, Kralj A, Young J, Meiser-Stedman R. The prevalence of intrusive memories in adult depression: A meta-analysis. J Affect Disord. 2019;253:193–202.

51. Lawrence-Wood E, Van Hooff M, Baur J, McFarlane AC. Re-experiencing phenomena following a disaster: The long-term predictive role of intrusion symptoms in the development of post-trauma depression and anxiety. J Affect Disord. 2016;190:278–281.

52. Lazarov A, Suarez-Jimenez B, Levi O, Coppersmith DDL, Lubin G, Pine DS, et al. Symptom structure of PTSD and co-morbid depressive symptoms - a network analysis of combat veteran patients. Psychol Med. 2020;50:2154–2170.

53. Koh J, Tng GYQ, Hartanto A. Potential and pitfalls of mobile mental health apps in traditional treatment: An umbrella review. Journal of Personalized Medicine. 2022;12:1376.

54. Abernethy A, Adams L, Barrett M, Bechtel C, Brennan P, Butte A, et al. The promise of digital health: Then, now, and the future. NAM Perspectives. 2022;2022.

55. Reading in the mobile era: a study of mobile reading in developing countries - UNESCO Digital Library. https://unesdoc.unesco.org/ark:/48223/pf0000227436. Accessed May 10, 2024.

56. The UN Specialized Agency for ICTs. Measuring digital development: Facts and Figures 2022. https://www.itu.int/hub/publication/d-ind-ict_mdd-2022/. Accessed May 10, 2024.

